# Common Genetic Variants Modify Disease Risk and Clinical Presentation in Monogenic Diabetes

**DOI:** 10.1101/2025.02.19.25322524

**Authors:** Jacques Murray Leech, Robin N Beaumont, Ankit M Arni, V. Kartik Chundru, Luke N Sharp, Kevin Colclough, Andrew T Hattersley, Michael N Weedon, Kashyap A Patel

## Abstract

The contribution of polygenic background in young onset monogenic disorders needs further exploration. Understanding this will provide new biological insights and may improve risk prediction of monogenic disease. Here we investigated the role of polygenic background in MODY—a common monogenic disorder with an age-dependent onset.We found strong enrichment of type 2 diabetes (T2D) polygenic risk, but not type 1 diabetes risk, in genetically confirmed MODY cases. This T2D polygenic burden, primarily through beta-cell dysfunction pathways, was strongly associated with earlier age of diagnosis and increased diabetes severity. Common genetic variants collectively accounted for 24% (p<0.0001) of the phenotypic variability. Using a large population cohort, we demonstrated that T2D polygenic burden substantially modifies diabetes onset in individuals with pathogenic variants, with diabetes risk ranging from 11% to 81%. Finally, we show that individuals with MODY-like phenotypes without a causal variant had elevated polygenic burden for T2D and related traits representing potential polygenic phenocopies. These findings reveal substantial influence of common genetic variation in shaping the clinical presentation of early-onset monogenic disorders. Incorporating these may improve risk estimates for individuals carrying pathogenic monogenic variants.

## Introduction

Growing evidence suggests that factors beyond the primary mutation play a greater role in rare monogenic conditions than previously recognised^1,2^. Monogenic diseases are classically defined by single causative mutations. Typically, these mutations are considered highly penetrant, with non-mutation factors such as polygenic and environmental background thought to play a small role, if any. However, large-scale population studies have revealed a more complex reality^3^. For example, our analysis of pathogenic *HNF1A* mutations that typically cause young-onset diabetes showed striking differences in diabetes penetrance: >90% in clinically ascertained cohorts versus <30% in population cohorts by age 40^4^. This pattern of unexpectedly low penetrance, now documented across multiple monogenic conditions^5,6^, suggests that polygenic and environmental factors may significantly influence disease manifestation, particularly in age-dependent monogenic conditions.

Maturity Onset Diabetes of the Young (MODY) serves as an excellent genetic disease model to investigate how common genetic variants influence young-onset monogenic disease. MODY is the most common autosomal dominant form of monogenic diabetes contributing up to 3% of all diabetes under age 30^7^. In this study we focused on the *HNF1A*, *HNF4A* and *HNF1B* genes (collectively referred to as HNF-MODY). The pathogenic variants in these three genes account for >90% of MODY cases^8–10^. These variants cause beta cell dysfunction leading to age-dependent diabetes typically presenting before age 25^9^. The availability of extensive genome-wide association data for both type 1 and type 2 diabetes and related metabolic traits, widely measured diabetes markers such as HbA1c allowing accurate diagnosis, and the availability of large MODY patient cohorts make MODY particularly suitable for studying common and rare disease interplay. Together, these resources provide a robust framework for examining how polygenic factors interact with young onset monogenic disorders.

Understanding these interactions is crucial both biologically and clinically. It can uncover new biological pathways and enhance disease prediction, knowledge that is essential for family counselling. This becomes increasingly important as genomic screening extends to clinically unselected cases and healthy newborns^11^. Previous studies have demonstrated that polygenic background can modify the penetrance of various monogenic conditions, including familial hypercholesterolaemia, obesity, kidney disease and Long QT syndrome^12–15^. These studies are important but often lack a defined age of disease onset. Initial studies of MODY suggested that Type 2 diabetes polygenic scores can influence age of diagnosis. However, these studies were limited by small sample sizes (<410) and focused solely on Type 2 diabetes variants ^16,17^, leaving the impact of Type 1 diabetes and other metabolic trait variants unexplored.

In this study, we investigated the interplay between polygenic background and age-dependent monogenic disorders, using MODY (maturity-onset diabetes of the young) as a model disease. In the largest MODY cohort studied to date, we demonstrate that common genetic variants explain a substantial proportion of phenotypic variation, disease expression and may explain MODY-like phenotypes.

## Results

### Polygenic burden of Type 2 diabetes is significantly enriched in genetically confirmed MODY

Our current understanding is that single rare pathogenic variant primarily drives the early onset diabetes of HNF-MODY, with limited contribution from common genetic background. We investigated this assumption by analysing polygenic scores (PGS) for type 2 diabetes (T2D), type 1 diabetes (T1D) and related metabolic traits (n=9) in 1,462 clinically referred patients with HNF-MODY (Tables S1, S2, S3). We compared these scores with those of 7,645 individuals without diabetes and 4,773 individuals with T2D (Table S1). We found significantly higher polygenic scores for T2D, Fasting Glucose, Fasting Insulin and Waist Hip Ratio in HNF-MODY patients compared to non-diabetic controls (0.09 - 0.42 standard deviation (SD) increase, all *P*<0.005) but no enrichment for T1D PGS (Figure 1A). The T2D PGS remained the strongest contributor (OR 1.46, 95% CI 1.36-1.58, *P*<0.0001) after accounting for other PGSs (Figure 1B). This enrichment was lower than observed in T2D cases and independent of parental diabetes history (Figure 1C) and after removing variants in *HNF1A*, *HNF4A* or *HNF1B* genes from the PGS (0.4 SD higher than control, 95% CI 0.35–0.46, *P*<0.0001). To identify which T2D pathways contributed to this enrichment, we analysed eight recently developed T2D pathway-specific hard cluster PGSs (Figure 1D). Of these, the metabolic syndrome (0.20 SD increase), residual glycaemic (0.17 SD) and beta-cell proinsulin positive (0.16 SD) pathway scores showed the strongest enrichments in MODY patients (all *P*<4×10^-8^). Sensitivity analyses by each gene and limited to probands showed consistent findings (Figures S1 and S2). Together, these data suggest that common T2D-associated variants contribute substantially to clinically diagnosed HNF-MODY.

**Figure 1:**
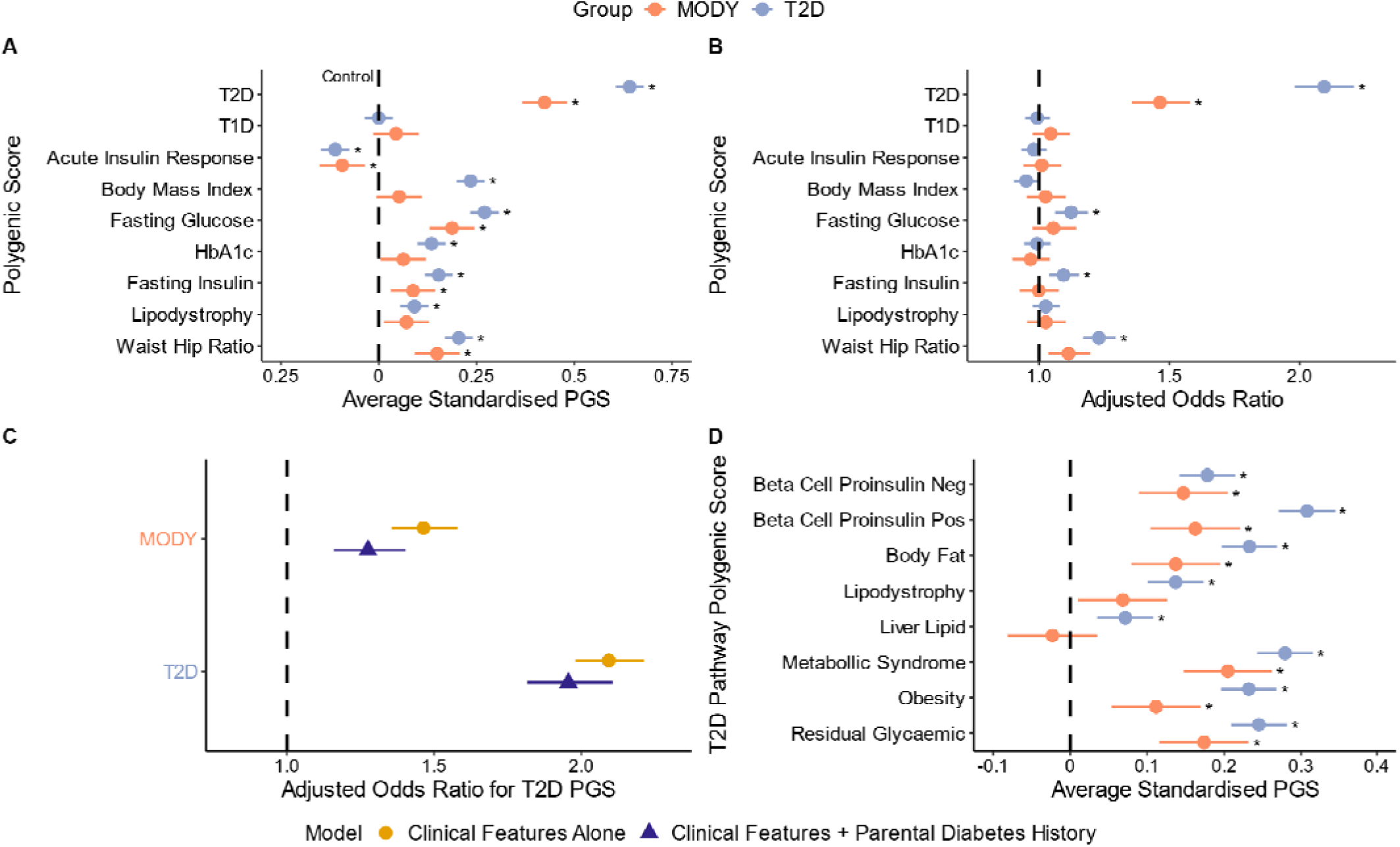
Elevated Polygenic Risk in HNF Monogenic Diabetes. **(A)** Standardised differences in upper-level diabetes-related polygenic scores, determined by linear regression adjusting for the first ten within-cohort principal components. HNF MODY carriers (orange, N=1,462) and T2D cases (blue, N=4,773) are compared to controls (dashed black line, N=7,645). **(B)** Adjusted odds ratios for T2D and HNF MODY cases versus controls, assessed using a logistic regression model including each PGS, sex, age, BMI, and the first ten within-cohort principal components as covariates. **(C)** Adjusted odds ratios for T2D PGS enrichment in HNF MODY and T2D cases under two models: (1) Adjusting for covariates as described in (B) (yellow) and (2) adjusting for the same covariates plus family history of diabetes (blue). **(D)** Standardised differences in T2D hard cluster partitioned polygenic scores. All scores are standardised to have a mean of 0 and standard deviation of 1 in controls. Odds ratios represent the change in risk associated with a 1 standard deviation increase in the respective polygenic score. Error bars represent 95% confidence intervals. Asterisks denote Bonferroni-adjusted statistically significant differences from controls.

### Increased type 2 diabetes polygenic burden was associated with an earlier onset and greater phenotypic severity in genetically confirmed MODY patients

We next assessed how the polygenic burden of T2D, T1D, and related metabolic traits influenced both the age of diagnosis and severity of diabetes in patients with clinically identified HNF-MODY. We defined diabetes severity as either requiring insulin treatment or having HbA1c ≥ 8.5% as proposed previously^18^. Only T2D PGS demonstrated a significant association with age of diagnosis after adjusting for other PGSs (*P*<3.3×10^-5^), with 1SD increase linked with a 1.19 years (0.68-1.7) earlier diagnosis (Figure 2A). In contrast, both the T2D and BMI PGSs were significantly associated with diabetes severity, with odds ratios of 1.24 (95% CI: 1.07–1.44, P = 0.004) and 1.32 (95% CI: 1.16–1.51, P < 3.1×10⁻□), respectively (Figure 2B). Our pathway analysis revealed that the beta cell proinsulin-positive pathway primarily drove the T2D PGS effect on diagnosis age (0.81 years [0.32-1.3] vs 0.67 0.15-1.18] years for all others combined) (Figure 2C). Whereas the obesity pathway demonstrated the strongest association with diabetes severity (OR 1.36, 1.19-1.56 vs 1.19,1.04-1.35 for all other pathways combined) (Figure 2D). Age of diagnosis and severity associations were maintained even after adjusting for clinical features. However, we observed strong effects of sex (females diagnosed 2.28 years earlier), maternal diabetes history (diagnosed 3.54 years earlier), and BMI (0.24-year earlier diagnosis) on age of diagnosis (Table S4, S5). Sensitivity analyses by each gene show directional consistent results (Table S6). These findings highlight the complex interaction between genetic and clinical factors that shape the clinical presentation of HNF-MODY.

**Figure 2:**
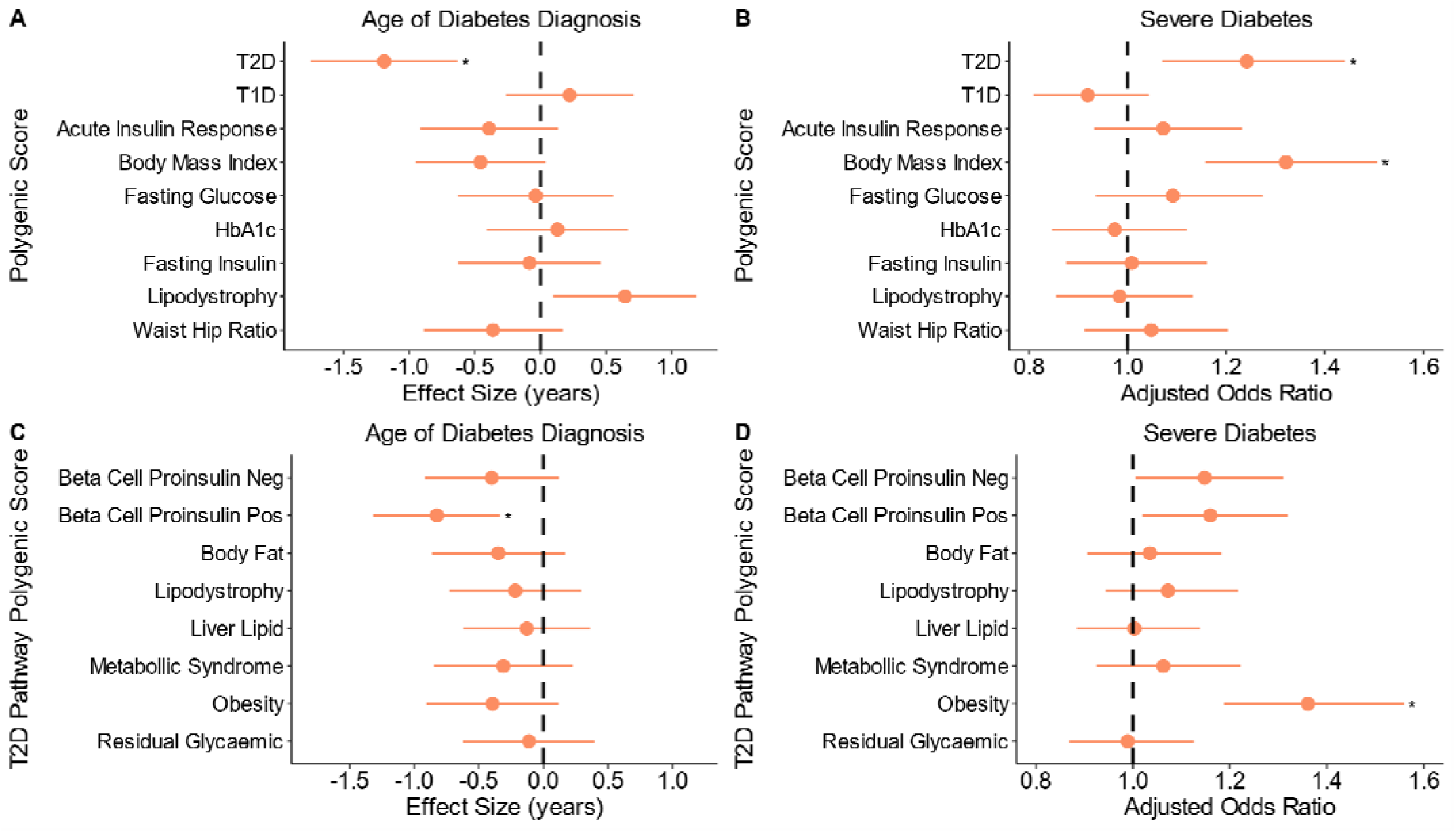
Increased Polygenic Risk Associated with earlier and more severe diabetes diagnosis in HNF MODY. **(A)** Association between polygenic scores for upper-level diabetes related traits and age of diabetes diagnosis. Estimates were derived using a mixed-effects linear model with family as a random effect and adjusted for other polygenic scores and the first ten within-cohort principal components. **(B)** Association between polygenic scores and risk of severe diabetes (defined as HbA1c ≥ 8.5% or insulin treatment at recruitment), using a mixed-effects logistic model with the same covariates as in (A). In total, 676 out of 1462 MODY carriers met the criteria for severe diabetes. Association of T2D-partitioned risk scores with age of diabetes diagnosis **(C)** and diabetes severity **(D)**, estimated using linear mixed effects models adjusted for the first ten within cohort principal components. Dots represent the estimates, with lines indicating 95% confidence intervals. Asterisks highlight significant differences after Bonferroni correction. Estimates represent the effect of a 1 standard deviation increase in the respective polygenic score.

### Type 2 diabetes polygenic burden modifies the risk of diabetes in clinically unselected carriers of pathogenic HNF-MODY variants

We next investigated how polygenic T2D background influences diabetes risk in carriers of pathogenic HNF-MODY variants. To assess this, we needed to investigate individuals not ascertained clinically to see the clear effect of polygenic background. Therefore, we analysed 424,553 European individuals with whole exome sequencing data from the clinically unselected UK Biobank population cohort. Among these, 100 individuals were identified as carriers of pathogenic variants in *HNF1A* (n= 34), *HNF4A* (n=51), or *HNF1B* (n=15) (Table S7, S8). Using a T2D PGS that did not include UK Biobank in the discovery cohort^19^, we found that among mutation carriers, diabetes risk varied substantially by T2D PGS. Compared to non-carriers with intermediate T2D PGS (middle three quintiles), carriers’ risk ranged from 8.5-fold (95% CI: 3.65-19.85) in those in the lowest T2D PGS quintile to 40.22-fold (95% CI: 14.95-108.24) in those in the highest quintile (Figure 3A). Despite the limited sample size, diabetes risk seemed to rise consistently across the range of T2D PGS, with diabetes risk ranging from 11.4% (1st percentile, 95% CI: 6.96-15.88) to 81.7% (99th percentile, 95% CI: 75.17-88.34) (Figure 3B). Interestingly, non-carriers in the 99th T2D PGS percentile showed a 17.7% risk (95% CI: 17.3-18.2) which was similar to mutation carriers with lowest T2D PGS. A sensitivity analysis using T2D PGS, which excluded variants within 1 MB of the three MODY genes, showed similar effect sizes (OR 2.17, 1.2-3.91 per 1SD change whole PGS vs 2.06, 1.16 – 3.82 without MODY genes). These data together suggest a significant contribution of T2D polygenic background on diabetes risk in HNF-MODY. While some individuals without MODY mutations but with extreme polygenic risk may reach to similar risk as HNF-MODY.

**Figure 3:**
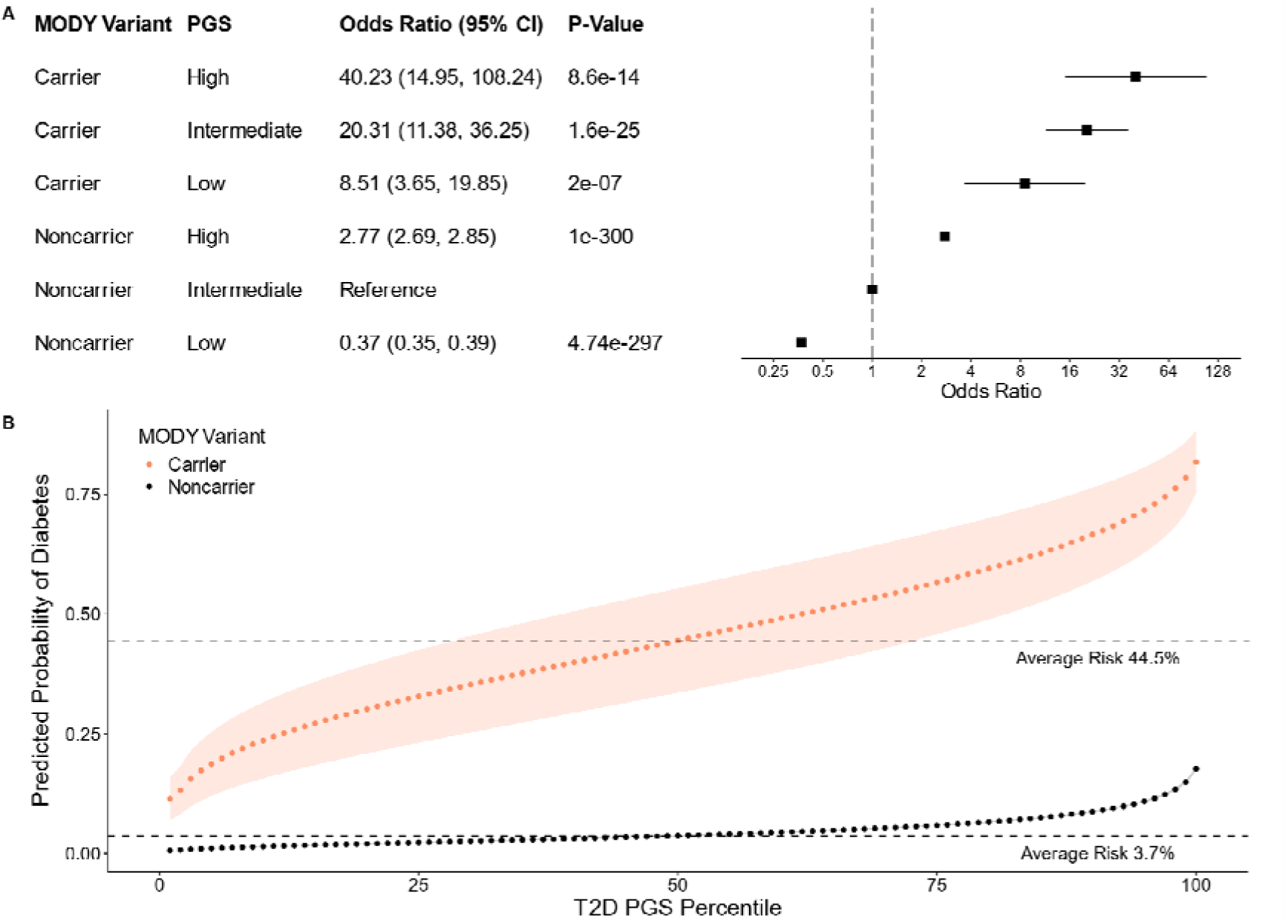
Polygenic background modifies Diabetes Risk in clinically unselected HNF MODY carriers. **(A)** Odds Ratios for diabetes risk in HNF-MODY carriers and non-carriers, stratified by T2D polygenic risk levels. Low risk is defined as the bottom 20% of T2D polygenic scores, high risk as the top 20%, and intermediate risk as the remaining 60%. N = 88,905, 265,079, and 70,469 for non-carriers, and N = 28, 55, and 17 for carriers, for low, intermediate, and high T2D PGS risk, respectively. Diabetes risk was estimated using a logistic regression model adjusted for sex, age, family history of diabetes, the first ten ancestry principal components, and BMI. **(B)** Predicted probability of diabetes at baseline across each percentile of T2D polygenic risk, assessed using a logistic model with T2D polygenic score as a continuous variable and the same covariates as in (A). Points represent percentiles, and shaded regions represent 95% confidence intervals. Dashed lines indicate the baseline diabetes risk at the 50th percentile of T2D polygenic scores for HNF-MODY carriers and non-carriers.

### Common genetic variants explain 24% of phenotypic variance in MODY

Having observed substantial contribution of polygenic background, we next aimed to quantify the overall contribution of common genetic variants to MODY. Using GCTA-GREML, we estimated common variant (MAF>0.01) SNP-heritability (h^2^), on the liability scale. Our analysis revealed a SNP heritability of 23.9% (95% CI: 17.2%-30.7%, p<0.0001) in individuals with HNF-MODY (Figure 4). This estimate was only slightly lower than in polygenic T2D cases 30.8% (95% CI: 25.08%-36.61%, p<0.0001). The heritability estimate remained consistent across multiple approaches, including restricting to HNF1A-related monogenic diabetes, phenotype-correlation genotype-correlation regression, and applying GREML estimation in LDAK (Table S9). To determine how much of this common variant heritability stems from T2D-associated variants, we calculated SNP-heritability for MODY comparing against 4,461 T2D cases, both with and without T2D PGS adjustment. The heritability decreased to 20.3% when compared against T2D, and further dropped to 17.2% (95% CI: 4.7%-29.7%, p = 0.035) after T2D PGS adjustment (Figure 4). These findings reveal that common genetic variants substantially influence MODY’s clinical presentation. At least one-third of this influence comes from T2D variants, suggesting the presence of T2D-independent genetic modifiers in HNF-MODY.

**Figure 4:**
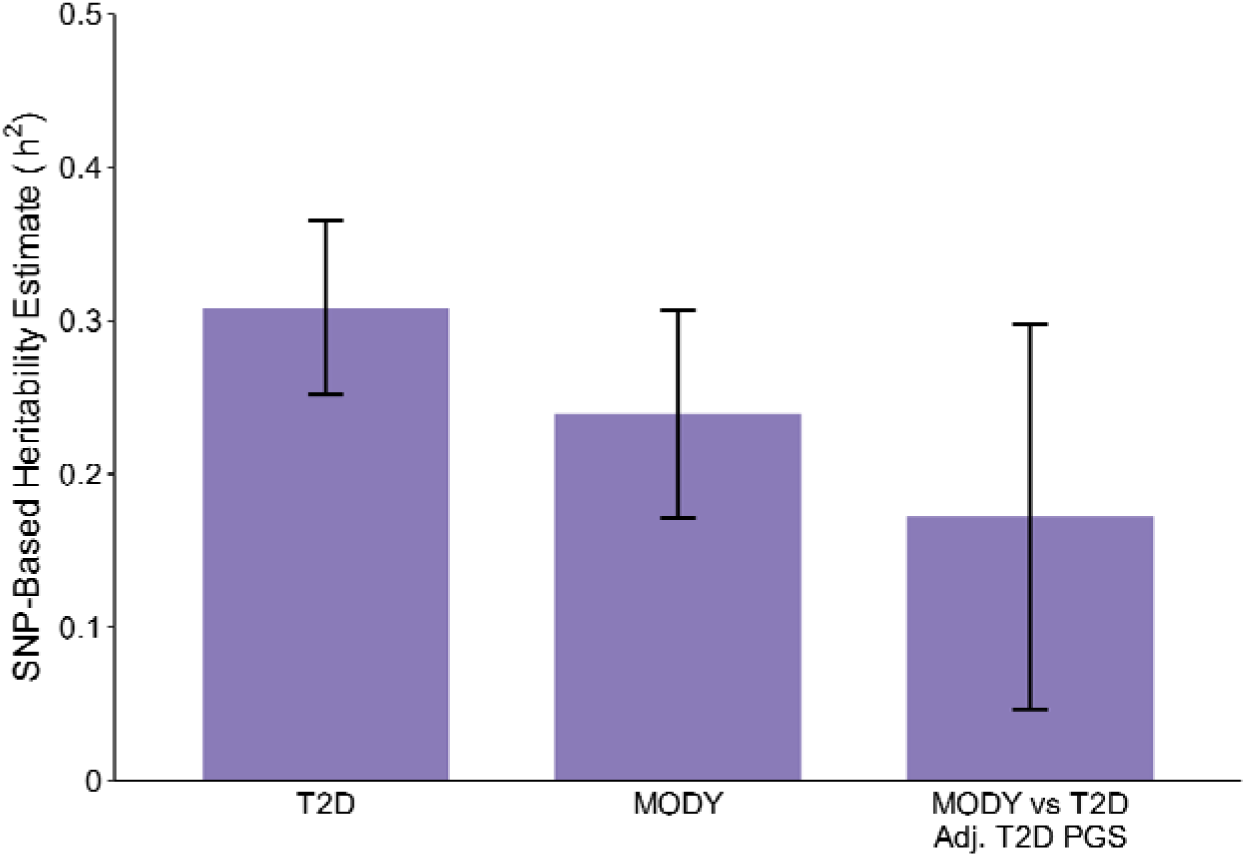
SNP-based heritability estimates for MODY. SNP-based heritability (h²) estimates were calculated in unrelated individuals using GCTA GREML-LDMS, stratified into four LD bins of equal size to construct the genetic relationship matrix, with sex, age, and the first 10 principal components within the cohort as covariates. Error bars represent the 95% confidence interval. Estimates for HNF-MODY carriers (N = 864) and T2D cases (N = 4,461) were compared to non-diabetic controls (N = 6,935). Heritability (h²) is shown on the liability scale for T2D (prevalence = 0.1) and MODY (prevalence = 0.0005). MODY and T2D cases were compared, adjusting for T2D polygenic score.

### Clinically referred MODY cases without a pathogenic variant have substantially higher polygenic burden of T2D and related traits

Following our observations of substantial common genetic variant contributions in mutation-positive MODY patients, we investigated whether higher polygenic background could also explain diabetes in individuals with a MODY phenotype but without causative mutations in known monogenic diabetes genes. We studied 300 individuals referred for MODY genetic testing from routine clinical practice with diabetes diagnosis before age 30 and BMI <30 kg/m², and without evidence of T1D (absence of positive islet autoantibodies or low C-peptide, high T1D GRS >50th centile of T1D population)^20^. These Unsolved MODY cases showed similar age of diagnosis and BMI to mutation-positive MODY cases (*p* > 0.05 for both) (Figure 5A, 5B, Table S10). As expected, these unsolved cases showed no excess T1D PGS but displayed a striking 1.18 SD (95% CI 1.07-1.29, p < 0.0001) higher T2D PGS than controls (Figure 5C). This polygenic burden was higher than both mutation-positive MODY cases by 0.73 SD and T2D cases by 0.52 SD (all p < 0.0001) (Figure S3). Compared to controls, we also observed an excess polygenic burden of BMI and Waist Hip Ratio in these cases (Figure 5D, 5E). Unsolved cases demonstrated an enrichment in all T2D partitioned PGSs, with the largest difference from controls in the beta cell proinsulin positive cluster (0.62 SD increase, 95% CI: 0.51 – 0.74, P < 0.0001) (Figure S4). Excess biparental diabetes history further supported the observed excess polygenic enrichment in Unsolved cases compared to T2D (53% one parent, 15.7% both parents with diabetes vs 28.9% and 4%, respectively in T2D) (Figure S5). These findings suggest that while some unsolved cases may harbour novel monogenic diabetes mutations, many likely represent polygenic phenocopies driven by an excessive polygenic burden of T2D and related traits.

**Figure 5:**
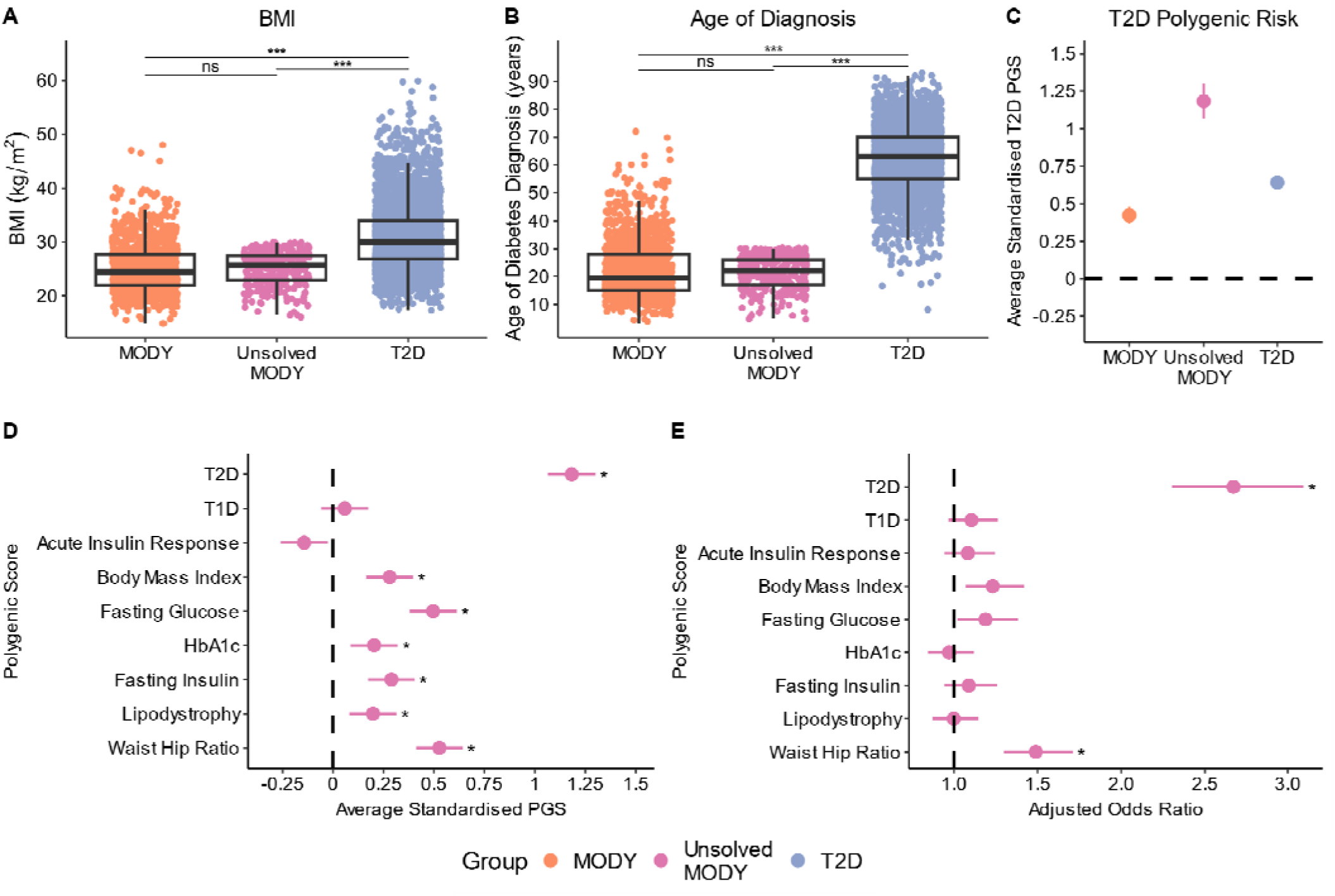
Unsolved MODY cases exhibit extreme T2D polygenic risk while phenotypically resembling typical MODY cases. Distribution of key characteristics and polygenic risk across MODY carriers (orange, n = 1462), T2D cases (blue, n = 4773), and Unsolved MODY cases (pink, n = 300). Distribution of clinical characteristics (**A)** Body Mass Index (BMI) and **(B)** Age of diabetes diagnosis, which were collected at patient referral. Box plots display the median and interquartile range, with individual data points overlaid. Statistical significance between groups is indicated by three asterisks (*** = P < 0.017, ns = nonsignificant.), as determined by t-tests. **(C)** Mean T2D polygenic risk across groups versus controls, assessed using a linear regression model adjusting for first ten within cohort principal components. **(D)** Mean polygenic risk score differences for diabetes-related traits in Unsolved MODY cases versus controls, using the same method as (C). **(E)** Adjusted odds ratios for Unsolved MODY cases versus controls, assessed using a logistic regression model including each PGS, sex, age, BMI, and the first ten within-cohort principal components as covariates. In (D) and (E), Asterisks denote significant differences from controls (P< 0.005). Dots represent point estimates, with error bars representing the 95% confidence intervals, in (C), (D) and (E).

## Discussion

In this study, we demonstrate that HNF-MODY has a significant polygenic component, with common genetic variation substantially influencing disease onset and severity in genetically confirmed MODY cases. The elevated polygenic burden of T2D-related traits may explain MODY phenocopies lacking pathogenic mutations.

MODY’s genetic architecture appears more complex than its traditional characterization as a purely monogenic disorder. We found that common genetic variations explain approximately 24% of phenotypic variance in clinically identified cases. This estimate is substantially higher than previously reported in other monogenic disorders (long QT syndrome-15%^15^, developmental delay-11%^21^) and approaches that of T2D. Such high polygenic contribution is unexpected for a presumed monogenic disease and may reflect its young-onset nature. Among analysed traits, type 2 diabetes (T2D) variants had the largest impact, likely due to overlapping genetic pathways. The observation aligns with previous small studies in HNF1A-MODY and studies in intermediate-effect MODY variants^16,17,19^. The absence of interaction with type 1 diabetes (T1D) polygenic risk aligns with the current understanding that T1D variants primarily affect autoimmune pathways rather than transcriptional networks disrupted in HNF-MODY^22^. This genetic distinction supports using T1D polygenic risk scores to differentiate MODY from early-onset T1D^20,23^.

Our findings reveal distinct genetic pathways modifying different aspects of MODY. Beta cell proinsulin-related variants predominantly influence age of diagnosis, while obesity-associated variants and beta cell pathways drive disease progression. This supports a liability threshold model where pathogenic MODY variants drive early-onset disease, with the polygenic background modifying overall disease risk. Notably, carriers with low T2D polygenic risk show substantially lower diabetes risk, with about half remaining disease-free in the population cohort. This explains the disparity between MODY prevalence in clinical referrals (1:10,000) versus genetic screening (1:2,000)^4,24^. Together, these data demonstrate that MODY’s pathogenesis involves substantial polygenic interaction rather than following a simple deterministic monogenic model.

Some unsolved MODY cases may represent polygenic phenocopies. Our small cohort of mutation-negative cases shows substantial enrichment of T2D polygenic risk exceeding that seen in typical T2D. This enrichment extends beyond T2D to other related traits, supporting complex polygenic aetiology. Additional factors such as intermediate-effect variants likely contribute to these MODY-like phenotypes, similar to young-onset obese T2D^25^. Our sample size precluded identification of specific pathway clusters, these cases may group into distinct pathways as observed in typical T2D^26^. Similar patterns are observed in other monogenic conditions like Long QT syndrome^15^, developmental delay^21^, and familial hypercholesterolemia^12^, where mutation-negative patients show higher polygenic burden than mutation-positive cases. Collectively, these findings suggest the presence of polygenic phenocopies. However, due to the relatively small sample size, these results should be considered preliminary. Further studies are needed to replicate these observations and elucidate the underlying mechanisms.

Our findings support the hypothesis that monogenic disorders exist on a continuum, where both pathogenic mutations and polygenic background shape disease manifestation^27^. Age-dependent conditions, such as MODY, are likely to have a larger polygenic contribution compared to neonatal-onset disorders. As evidence accumulates, this observation may extend to the majority of monogenic disorders, albeit to varying degrees. However, each condition will require individual evaluation to quantify the relative contributions. With the declining cost of genetic testing and the increasing identification of presymptomatic carriers through incidental findings^28^ and newborn screening programs^29^, there is a growing need to refine disease risk prediction. Currently, risk assessment relies solely on the presence of pathogenic mutations. To provide more precise risk stratification, it may be necessary to incorporate non-mutation factors, such as polygenic risk scores or family history, as is already done in conditions like breast cancer^30^. As we move toward whole-genome sequencing as a first-line test, a single assessment could provide comprehensive genetic information, including both monogenic and polygenic risk factors, if desired. However, further studies are needed to evaluate the clinical utility of this approach in improving disease prediction.

Although this represents the largest MODY study to date, several limitations warrant consideration. Our sample size was limited for individual MODY genes and unsolved cases, constraining gene-specific analyses. The predominantly UK-based, European ancestry cohort limits generalizability to other populations. While MODY variants in the UK Biobank weren’t Sanger-confirmed, we minimised false positives through manual IGV review and strict quality filtering. The UK Biobank’s healthy volunteer bias likely underestimates true MODY penetrance in general populations due to underrepresentation of early-onset diabetes. Furthermore, our sample size restricted detailed pathway analysis of unsolved MODY cases.

In summary, using MODY as a model disease, we demonstrate substantial interplay between monogenic mutations and polygenic background in young-onset monogenic disorders. Our findings suggest that future approaches to disease prediction will require integration of monogenic, polygenic, and environmental factors to improve clinical utility.

## METHODS

### Study Populations

#### Exeter MODY Cohort

##### MODY Individuals with Confirmed Pathogenic Variants

We analysed individuals referred for monogenic diabetes genetic testing at the Exeter Genomics Laboratory, Royal Devon University Healthcare NHS Foundation Trust, Exeter, UK. These referrals originated from clinical suspicion of MODY during routine clinical care in the UK. These individuals were found to have likely pathogenic or pathogenic variants either by Sanger sequencing or gene panel test performed as part of routine clinical care. Our cohort comprised European individuals with diabetes and carrying pathogenic variants in HNF1A (n = 997), HNF1B (n = 145), or HNF4A (n = 320).

##### Unsolved MODY Individuals

We evaluated 300 European individuals referred from routine clinical care in the UK with suspected MODY. All participants received their diabetes diagnosis before age 30 and lacked clinical features suggestive of typical type 2 diabetes (BMI ≥30 kg/m²) or type 1 diabetes (positive islet autoantibodies, C-peptide <200 pmol/L, and a ten-SNP T1D genetic risk score above the 50th centile of the gold-standard T1D population from the WTCCC study)^20^. These individuals underwent comprehensive genetic testing for all known monogenic diabetes genes (n = 58) and were not found to have pathogenic variants in these genes. All probands or their guardians provided informed consent, and the North Wales Ethics Committee approved the study. The clinical features of these solved and unsolved MODY cases, at referral for genetic testing, are summarized in Table S1.

#### Type 2 Diabetes and Non-Diabetes Control Cohort

We analysed participants from two ethically approved population cohorts in Southwest England: The Exeter 10000 study (https://exetercrfnihr.org/about/exeter-10000/)^31^ and The Diabetes Alliance for Research in England (DARE) Study (https://www.diabetesgenes.org/current-research/dare/)^32^. These studies recruited unselected participants through primary care practices across Southwest United Kingdom. At recruitment, participants completed baseline questionnaires and provided fasting blood and urine samples for measurement of diabetes-related markers, including fasting glucose and HbA1c. Our analysis included European individuals who underwent array genotyping as part of these studies. We classified participants as having type 2 diabetes if they either did not require insulin treatment or initiated insulin treatment after 36 months from diagnosis, thereby excluding potential misclassified type 1 diabetes cases. We defined controls as individuals without a known diabetes diagnosis and HbA1c ≤48 mmol/mol (6.5%)^33^. The final cohort comprised 7,645 controls and 4,773 individuals with type 2 diabetes, with their clinical characteristics presented in Table S1. The NIHR Exeter Clinical Research Facility management committee approved access to these samples and genotype data.

#### UK Biobank Cohort

The UK Biobank represents a large-scale, prospective population-based study comprising approximately 500,000 UK residents aged 40-70 years at enrolment^34^. Recruitment occurred between 2006 and 2010, with comprehensive data collection through multiple channels: participant questionnaires, structured interviews, and biomarker measurements^34^. The study supplemented this information with medical history data from HES records coded using ICD-9 and ICD-10 codes. We defined diabetes status using three criteria: self-reported diagnosis, HbA1c levels ≥6.5 % at recruitment, or active diabetes treatment at recruitment. Our study cohort compromised 424,553 European individuals who underwent exome sequencing and array genotyping. Clinical characteristics of these individuals can be found Table S7. We analysed the exome sequence data to identify individual with likely pathogenic and pathogenic variants in *HNF1A/HNF4A/HNF1B* as described before^4^, with details of variants identified in Table S8.

#### Genetic Analysis

##### MODY Pathogenic Variants in Exeter MODY Cohort and UK Biobank

For the Exeter MODY cohort, all referred patients were screened for potential MODY-associated variants using either Sanger sequencing or gene panel testing, following methodologies detailed in Ellard *et al*^35^. For the UK Biobank participants, we utilised exome sequence data to identify carriers of pathogenic MODY variants. We annotated all variants using clinically validated transcripts: GenBank NM_000545.6 for *HNF1A*, NM_000458.4 for *HNF1B*, and NM_175914.4 for *HNF4A*. We classified variants according to the American College of Medical Genetics and Genomics (ACMG)/Association of Molecular Pathology (AMP) guidelines, designating them as either likely pathogenic (class 4) or pathogenic (class 5)^36^. This classification process followed our established protocols for the local Exeter cohort and aligned with our recent study’s methodology^4^. Table S8 presents a comprehensive list of variants identified in the UK Biobank cohort.

#### Array Genotyping

##### Exeter MODY, T2D and Non-Diabetic Controls

We performed array genotyping using the Infinium Global Screening Array platform. Our comprehensive quality control protocol excluded samples with call rates below 98%, sex mismatches, relationship discrepancies, or inbreeding coefficients exceeding 0.1. At the variant level, we removed markers with missingness above 2%, minor allele frequency below 5%, or deviation from Hardy-Weinberg equilibrium (p < 1e-6). We applied these quality control measures both independently for each batch and following batch integration. We then used linkage disequilibrium (LD) pruned markers for genotype imputation through the TOPMed reference panel v2^37^ via the Michigan Imputation Server^38^. To determine genetic ancestry, we compared our data against reference populations from the 1000 Genomes Phase 3 and Human Genome Diversity Project, implementing the principal components analysis approach within the GenoPred Pipeline (v2.2.1)^39,40^. For relationship inference, we analysed LD pruned data using the KING robust algorithm (v2.2.4) to identify unrelated individuals up to the third degree^41^. To better capture the within-cohort population structure, we conducted principal component analysis using FlashPCA (v2.0)^42^. Initially, we calculated principal components in unrelated European individuals and then projected these onto related European individuals.

##### UK Biobank

The UK Biobank individuals were SNP-genotyped using the UK BiLEVE array for the first ∼50,000 individuals, with the remaining using the UKB Axiom array. This dataset underwent central quality control by the UK Biobank and was imputed to the TOPMed reference panel^37^. Approximately 450,000 individuals from the UKB Array also underwent exome sequencing using the IDT xGen Exome Research Panel v1.0. Detailed sequencing methodology for UK Biobank samples has been described previously^43^. In brief, variants were called using GATK 3.0 filtering variants with an inbreeding coefficient < −0.03 or without at least one variant genotype of DPD≥D10, GQD≥D20 and, if heterozygous, ABD≥D0.20. For this analysis, we included 424,553 individuals who had both exome and array data and were of European ancestry, inferred from projected principal component analysis using the same approach as for the local cohort.

#### Polygenic Score Calculation

We calculated polygenic scores for T2D^44^, T1D^45^ and seven diabetes related traits^46–50^, alongside eight pathway-specific T2D risk scores^44^. We constructed weighted polygenic scores using genome-wide significant variants. For traits with comprehensive GWAS summary statistics available, we implemented genome-wide calculations to capture additional genetic signal. Our computational pipeline utilised PLINK 1.9’s score function for genome-wide significant variant-based scores^51^. For the genome-wide polygenic scores, we implemented the GenoPred 2.2.1 pipeline with LDpred2’s auto model, enabling comprehensive processing of GWAS summary statistics^40,52^. Further details are available in Table S3, including the specific approach used for each trait, including the calculation method, number of variants incorporated, and the source GWAS studies.

#### Heritability Estimation

To estimate the common variant contribution to MODY and T2D, SNP based heritability was estimated in unrelated individuals using GCTA GREML-LDMS, stratifying into four LD bins of equal size to construct the genetic relationship matrix.^53^ To test the validity of these estimates we ran phenotype correlation–genotype correlation and restricted maximum likelihood approaches implemented in LDAK, using thinned predictors to construct the kinship matrix^54,55^. We used sex, age and the first 10 within cohort PCs as covariates for each method. For MODY, disease prevalence was set at 0.0005^4^ and 0.00025^24^, and for T2D, at 0.1^56^ (Table S9). Variants with an imputation quality > 0.9 and minor allele frequency > 1% were used to in this analysis.

### Statistical Analysis

To assess polygenic risk in MODY carriers and T2D cases, we employed several different steps. Linear models were used to identify polygenic scores enriched in MODY carriers. We used logistic models, adjusting for clinical characteristics and other PGSs, were used to identify scores which could independently separate cases and controls. Unadjusted and adjusted linear and logistic mixed models were used to assess the relationship between PGS and age of diagnosis and diabetes severity, including family ID as a random effect to model within family effects. In the UK Biobank, individuals were stratified into low, intermediate or high PGS groups, defined by the bottom quintile, middle three quintiles or top quintile for T2D risk. Odds ratio of diabetes risk was calculated using a logistic regression model, using non MODY carriers of intermediate T2D risk as reference. To estimate the probability of diabetes across T2D percentile, we used a logistic model with T2D PGS as a continuous covariate alongside carrier status and clinical characteristics and computed the marginal effects per percentile. All statistical analysis carried out using R version 4.4.1 and Stata version 18.

## Author Contributions

Concept and Design: K.A.P, M.N.W, A.T.H, J.M

Acquisition, analysis, or interpretation of data: J.M, K.A.P, A.M.A, R.N.B, K.C, V.K.C, L.N.S

All authors contributed to revisions and the final manuscript.

K.A.P is the guarantor of the work.

The research utilised data from the UK Biobank resource carried out under UK Biobank application number 103356. UK Biobank protocols were approved by the National Research Ethics Service Committee. KAP is funded by Wellcome Trust (219606/Z/19/Z), ATH is supported by Wellcome Trust Senior Investigator award (WT098395/Z/12/Z). The work is supported by the National Institute for Health Research (NIHR) Exeter Biomedical Research Centre, Exeter, UK. The Wellcome Trust, MRC and NIHR had no role in the design and conduct of the study; collection, management, analysis, and interpretation of the data; preparation, review, or approval of the manuscript; and decision to submit the manuscript for publication. The views expressed are those of the author(s) and not necessarily those of the Wellcome Trust, Department of Health, NHS or NIHR. For the purpose of open access, the author has applied a CC BY public copyright licence to any Author Accepted Manuscript version arising from this submission.

## Data Availability

The data supporting the findings of this study are available within the article and its supplemental information. The clinical data generated and/or analysed as part of this study are not publicly available because of patient confidentiality and ethical approval associated with the data but are available from the corresponding authors upon reasonable request. The UK Biobank dataset is available from https://biobank.ctsu.ox.ac.uk

