## Supplementary Data 1 for "Common Genetic Variants Modify Disease Risk and Clinical Presentation in Monogenic Diabetes"

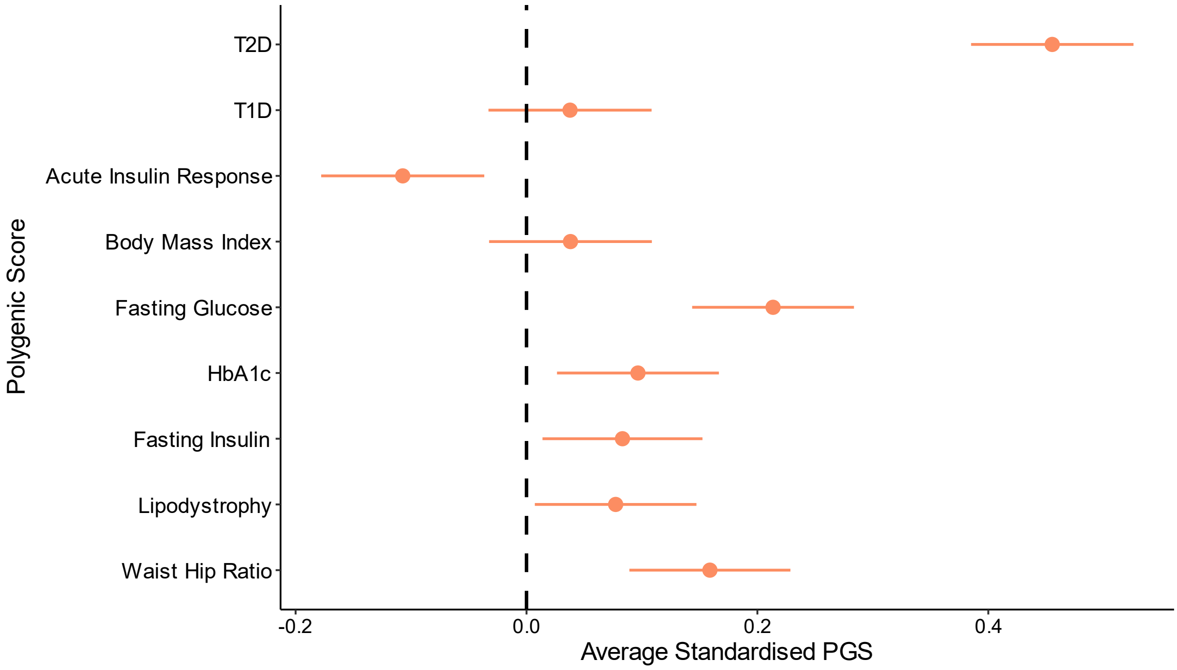


Figure S1: Polygenic Score difference from controls restricted to probands only.

Mean polygenic score difference for MODY probands (orange, N=924) compared to controls (dashed black line, N=7645). Scores are standardized, with controls set to a mean of 0 and a standard deviation of 1. Error bars indicate 95% confidence intervals, and dots represent the mean estimates, determined by linear regression models adjusted for the first ten within-cohort principal components.

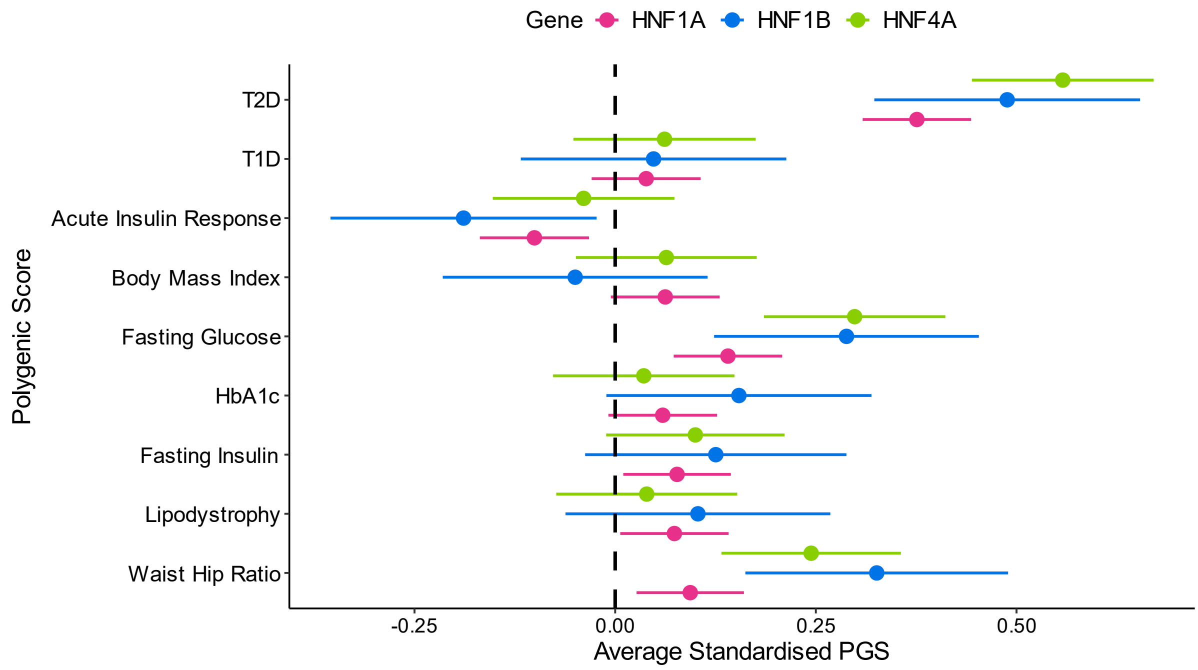


Figure S2: Polygenic Score difference from controls by HNF MODY subtype.

Mean polygenic score differences for HNF1A (pink, N=997), HNF1B (blue, N=145), and HNF4A (green, N=320) carriers compared to controls (dashed black line, N=7645), as determined by linear regression models adjusted for the first ten within-cohort principal components. Scores are standardized, with controls set to a mean of 0 and a standard deviation of 1. Error bars represent 95% confidence intervals, and dots denote the mean estimates.

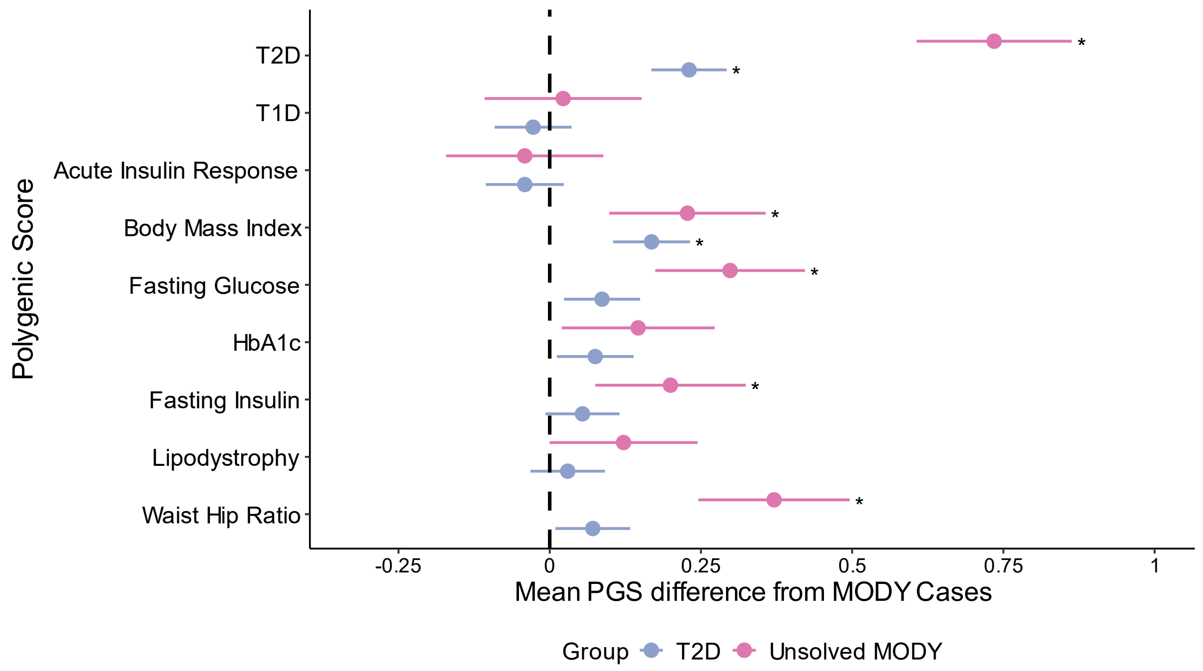


Figure S3: Mean polygenic score difference from solved MODY cases

Mean polygenic score difference between unsolved MODY cases (N = 300, pink) T2D cases (N = 4773, blue) versus MODY cases (N = 1462, assessed using a logistic regression model including the first ten within-cohort principal components as covariates. Asterisks denote significant differences (P<0.0028). Polygenic Scores were standardized so that the control population has a mean of 0 and standard deviation of 1. Error bars represent 95% confidence intervals.

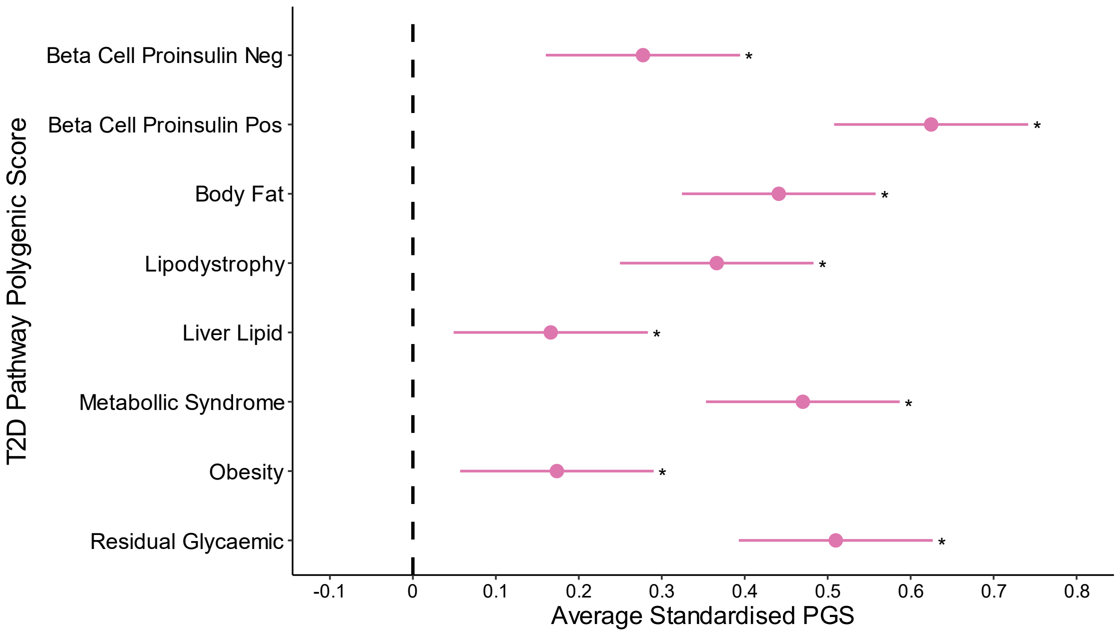


Figure S4: T2D Hard Cluster Polygenic Score Differences in Unsolved MODY Cases Compared to Controls

Mean polygenic score differences for T2D hard cluster partitioned scores in Unsolved MODY cases (pink, N = 300) compared to non-diabetic controls (dashed black line, N = 7,645). Scores are standardized with controls set to a mean of 0 and a standard deviation of 1. Error bars represent 95% confidence intervals, and dots indicate the mean estimates. Asterisks denote statistically significant differences from controls ($P < 0.006$), based on the Bonferroni significance threshold, as determined by linear regression models adjusted for the first ten within-cohort principal components.

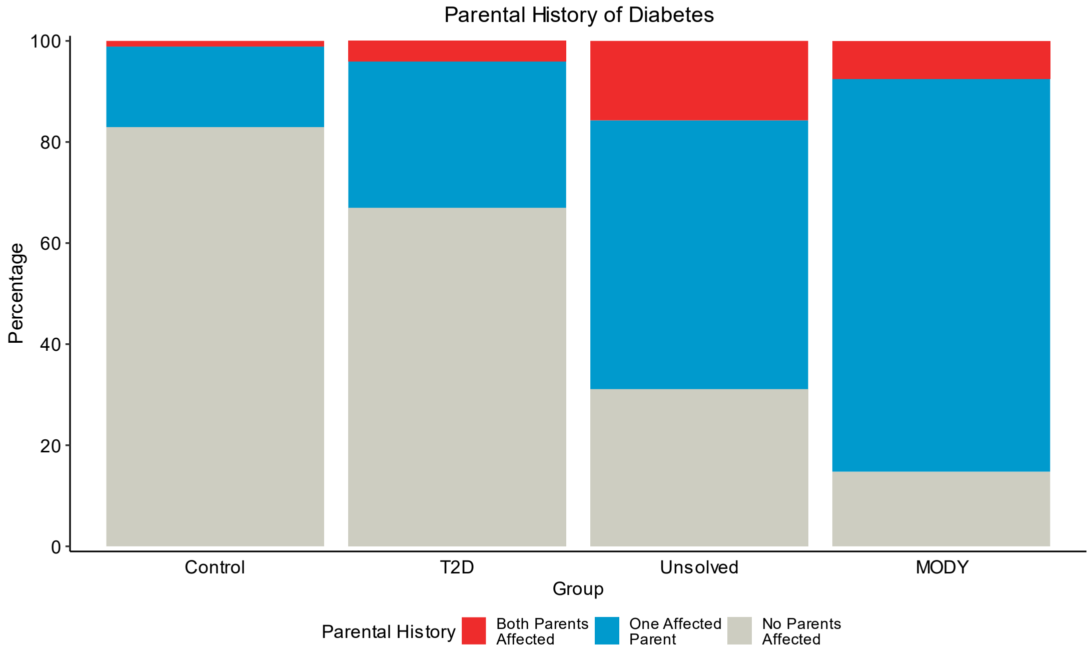


Figure S5: Unsolved MODY cases have a high percentage of familial diabetes history.

The plot shows the distribution of parental family history of diabetes for four groups: non-diabetic controls (N = 7,645), genetically confirmed MODY cases (N = 1,462), Unsolved MODY cases (N = 300), and type 2 diabetes cases (N = 4,773). Family history is categorized into three groups: no parent with diabetes (grey), one parent with diabetes (blue), and both parents with diabetes (red). Percentages are calculated within each group.

| Characteristics | Control | MODY (genetically confirmed mutation in *HNF1A/HNF4A/HNF1B*) | T2D | Unsolved MODY (monogenic diabetes gene panel negative) |
|---|---|---|---|---|
| N | 7,645 | 1,462 | 4,773 | 300 |
| Female Sex, n (%) | 4,849 (63.4) | 930 (63.6) | 1,957 (41) | 177 (59) |
| Diabetes, n (%) | 0 (0) | 1,462 (100) | 4,773 (100) | 300 (100) |
| Age at Recruitment, y | 54.3 (15.15) | 43.73 (18.29) | 68.28 (10.27) | 36.55 (12.45) |
| Age at Diabetes Diagnosis, y | - | 22.24 (9.87) | 61.99 (11.76) | 21.65 (6.34) |
| BMI (kg/m$^2$) | 26.48 (4.68) | 25.2 (4.63) | 30.87 (5.79) | 25.06 (3.14) |
| Parent Diabetes, n (%) | | | | |
| None | 6,205 (82.9) | 197 (14.8) | 1,203 (67) | 93 (31.1) |
| Mother | 571 (7.6) | 609 (45.7) | 277 (15.4) | 86 (28.8) |
| Father | 621 (8.3) | 427 (32) | 243 (13.5) | 73 (24.4) |
| Both | 84 (1.1) | 100 (7.5) | 73 (4.1) | 47 (15.7) |
| Insulin Treated, n (%) | 0 (0) | 539 (40) | 325 (11.3) | 68 (22.7) |
| HbA1c, % | 5.61 (0.4) | 7.64 (1.76) | 7.19 (1.12) | 8.01 (2.15) |

Table S1: Characteristics of participants in local cohort at referral for genetic testing. For continuous variables, values are presented as mean (SD), and for categorical variables, counts (n) and percentages (%) are provided. BMI = Body Mass Index, y = years.

| Characteristics | HNF1A | HNF1B | HNF4A |
|---|---|---|---|
| N | 997 | 145 | 320 |
| Female Sex, n (%) | 641 (64.3) | 76 (52.4) | 213 (66.6) |
| Diabetes, n (%) | 997 (100) | 145 (100) | 320 (100) |
| Age at Recruitment, y | 44.06 (18.67) | 36.63 (14.86) | 45.91 (17.85) |
| Age at Diabetes Diagnosis, y | 21.66 (9.47) | 22.06 (9.58) | 24.15 (10.94) |
| BMI ($kg/m^2$) | 25.01 (4.45) | 24.64 (5.18) | 25.94 (4.89) |
| Parent Diabetes, n (%) | | | |
| None | 101 (11) | 55 (48.7) | 41 (13.7) |
| Mother | 421 (45.7) | 37 (32.7) | 151 (50.5) |
| Father | 330 (35.8) | 16 (14.2) | 81 (27.1) |
| Both | 69 (7.5) | 5 (4.4) | 26 (8.7) |
| Insulin Treated, n (%) | 349 (37.6) | 76 (64.4) | 114 (38.0) |
| HbA1c, % | 7.56 (1.64) | 7.9 (2.44) | 7.73 (1.78) |

Table S2: Characteristics of clinically referred MODY cases by HNF subtype, at time of referral for genetic testing. For continuous variables, values are presented as mean (SD), and for categorical variables, counts (n) and percentages (%) are provided. BMI = Body Mass Index, y = years.

| Trait | PubMed ID | N SNPs in Score | Comments |
|---|---|---|---|
| Type 2 Diabetes (T2D) | 38374256 | 1289 | Constructed using plink – score function using genome wide significant variants |
| Type 2 Pathway Scores | 38374256 | 3 - 389 | Pathway specific scores of T2D |
| Type 1 Diabetes (T1D) | 30655379 | 67 | Weighted T1D score using T1DGRS2, available at: |
| Acute Insulin Response | 28490609 | 955764 | Genome-wide polygenic scores, we implemented the GenoPred 2.2.1 pipeline with LDpred2's auto model, which included quality control of summary statistics and genetic data |
| Body Mass Index (BMI) | 25673413 | 886707 | |
| Fasting Glucose | 34059833 | 1038695 | |
| Fasting Insulin | 34059833 | 1036765 | |
| HbA1c | 34059833 | 1039883 | |
| Waist Hip Ratio | 30239722 | 906879 | |
| Lipodystrophy | 27841877 | 53 | Constructed using plink – score function using genome wide significant variants |
| Type 2 Diabetes (T2D)* | 39379762 | 1087858 | Genome-wide polygenic score for T2D, using weights previously derived using PRS- CS, which excluding UKBB participants during testing |

Table S3: Polygenic Scores used in analysis. Polygenic scores (PGS) for various traits, including Type 1 and Type 2 Diabetes alongside other diabetes related traits, used in the analysis. For each trait, the corresponding PubMed ID, and the number of SNPs included in the score are provided. * Used in UK Biobank analysis as it does contain have weights from UK Biobank.

| Predictor | **Effect Size**<br>**(years earlier diabetes diagnosis)**<br>**(95% CI)** | ***P*** |
|---|---|---|
| T2D PGS (per SD increase) | 1.01 (0.49 – 1.54) | 1.54×10−4 |
| Sex (with respect to females) | 2.28 (1.17 – 3.39) | 5.56×10−5 |
| BMI (per $kg/m^2$ increase) | 0.24 (0.12 – 0.35) | 6.18×10−5 |
| Parent Diabetes History | | |
| Mother | 3.54 (1.89 – 5.17) | 2.55×10−5 |
| Father | 0.01 (-1.74 – 1.73) | 0.99 |
| Both | 0.01 (2.21 – 2.33) | 0.99 |

Table S4: T2D Polygenic lowers age of diagnosis in MODY cases even after adjusting for clinical characteristics. Upper-level polygenic scores identified as independently associated with age of diagnosis in Figure 2A were included in a mixed effect linear regression with age of diabetes as the outcome, and family ID as the random effect. Covariates included in the model: T2D PGS, Sex, BMI, Parental History, Gene, Variant Location, Year of Diabetes diagnosis, Proband/Family Member, and 10 within-cohort principal components. All effect sizes in years. Parental Diabetes in reference to subjects whose parents had no history of diabetes.

| Predictor | | Odds Ratio (Diabetes Severity) (95% CI) | *P* |
|---|---|---|---|
| T2D PGS (per SD increase) | | 1.23 (1.06 – 1.43) | 0.006 |
| Body Mass Index PGS (per SD increase) | | 1.31(1.13 -1.52) | 4.62×10−4 |
| Sex (with respect to females) | | 1.05 (0.77 – 1.44) | 0.74 |
| BMI (per $kg/m^2$ increase) | | 1.07 (1.04 – 1.11) | 9.54×10−5 |
| Parent Diabetes History | | | |
| | Mother | 1.38 (0.85 – 2.23) | 0.18 |
| | Father | 0.78 (0.47 – 1.29) | 0.32 |
| | Both | 1.63 (0.83 – 3.22) | 0.15 |

Table S5: T2D and BMI Polygenic increase diabetes severity in MODY cases, even after adjusting for clinical characteristics. Upper-level polygenic scores identified as independently associated with diabetes severity in Figure 2B were included in a mixed effect logistic regression with age of diabetes as the outcome, and family ID as the random effect. Covariates included in the model: T2D PGS, Body Mass Index PGS, Sex, BMI, Parental History, Gene, Variant Location, Year of Diabetes diagnosis, Proband/Family Member, and 10 within-cohort principal components. Parental Diabetes in reference to subjects whose parents had no history of diabetes. In total, 676 out of 1462 MODY carriers met the criteria for severe diabetes (defined as HbA1c ≥ 8.5% or insulin treatment at recruitment).

| Characteristics | | Noncarriers | MODY carriers |
|---|---|---|---|
| N | | 424,453 | 100 |
| Female Sex, n (%) | | 230,412 (54.28) | 59 (59%) |
| Diabetes, n (%) | | 24,371 (5.74) | 49 (49) |
| Age at Recruitment, y | | 57.26 (8.03) | 57.33 (7.68) |
| Age at Diabetes Diagnosis, y | | 59.45 (12.3) | 39.01 (17.38) |
| BMI ($kg/m^2$) | | 27.4 (4.75) | 26.15 (3.92) |
| Parent Diabetes, n (%) | | | |
| | None | 355,550 (83.77) | 53 (53) |
| | Mother | 33,069 (7.79) | 22 (22) |
| | Father | 31,717 (7.47) | 21 (21) |
| | Both | 4,117 (0.97) | 4 (4) |
| Insulin Treated, n (%) | | 4,385 (11.58) | 14 (27.5) |
| HbA1c, % | | 5.64 (0.56) | 6.39 (1.11) |

Table S7: Clinical Characteristics of UK Biobank Participants at Recruitment. Clinical characteristics of UK Biobank participants, split by MODY carriers and noncarriers, at the time of recruitment. For continuous variables, values are presented as mean (SD), and for categorical variables, counts (n) and percentages (%) are provided. BMI = Body Mass Index, y = years.

| Characteristics | MODY | Unsolved MODY | P Value |
|---|---|---|---|
| N | 1,462 | 300 | - |
| Female Sex, n (%) | 930 (63.6) | 177 (59) | 0.13 |
| Diabetes, n (%) | 1,462 (100) | 300 (100) | 1 |
| Age at Recruitment, y | 43.73 (18.29) | 36.55 (12.45) | $1.1 \times 10^{-10}$ |
| Age at Diabetes Diagnosis, y | 22.24 (9.87) | 21.65 (6.34) | 0.32 |
| BMI (kg/m$^2$) | 25.2 (4.63) | 25.06 (3.14) | 0.63 |
| Parent Diabetes, n (%) | | | $2.2 \times 10^{-16}$ |
| None | 197 (14.8) | 93 (31.1) | |
| Mother | 609 (45.7) | 86 (28.8) | |
| Father | 427 (32) | 73 (24.4) | |
| Both | 100 (7.5) | 47 (15.7) | |
| Insulin Treated, n (%) | 539 (40) | 68 (22.7) | $2 \times 10^{-8}$ |
| HbA1c, % | 7.64 (1.76) | 8.01 (2.15) | 0.0037 |

Table S10: Comparison of clinical features, collected at referral for genetic testing, between genotype-positive and genotype-negative (Unsolved) MODY cases. Continuous variables were assessed using t-tests, and categorical variables were assessed using chi-square tests. For continuous variables, values are presented as mean (SD). For categorical variables, counts (n) and percentages (%) are provided. BMI = Body Mass Index, y = years.
