## Supplementary Data 2 for "Common Genetic Variants Modify Disease Risk and Clinical Presentation in Monogenic Diabetes"

| PGS | Gene | Beta (Age Diagnosis) | Lower CI | Upper CI | P Value | Odds Ratio | Odds Ratio Lower CI | Odds Ratio Upper CI | P Value |
|---|---|---|---|---|---|---|---|---|---|
| T2D | HNF1A | -1.57 | -2.24 | -0.90 | 4.51E-06 | 1.27 | 1.04 | 1.56 | 2.21E-02 |
| T2D | HNF1B | -0.76 | -2.34 | 0.82 | 3.43E-01 | 1.31 | 0.57 | 3.04 | 5.25E-01 |
| T2D | HNF4A | -0.83 | -2.12 | 0.46 | 2.05E-01 | 1.09 | 0.78 | 1.53 | 6.17E-01 |
| T1D | HNF1A | 0.29 | -0.28 | 0.86 | 3.21E-01 | 0.95 | 0.80 | 1.12 | 5.41E-01 |
| T1D | HNF1B | -0.72 | -2.31 | 0.87 | 3.73E-01 | 2.38 | 0.80 | 7.08 | 1.19E-01 |
| T1D | HNF4A | 0.12 | -0.98 | 1.21 | 8.32E-01 | 1.07 | 0.80 | 1.44 | 6.30E-01 |
| Acute Insulin Response | HNF1A | -0.59 | -1.21 | 0.02 | 5.94E-02 | 1.03 | 0.84 | 1.24 | 8.01E-01 |
| Acute Insulin Response | HNF1B | -1.87 | -3.51 | -0.23 | 2.60E-02 | 1.16 | 0.36 | 3.71 | 8.07E-01 |
| Acute Insulin Response | HNF4A | 0.60 | -0.67 | 1.86 | 3.53E-01 | 1.31 | 0.92 | 1.87 | 1.34E-01 |
| Body Mass Index | HNF1A | -0.41 | -0.98 | 0.17 | 1.64E-01 | 1.30 | 1.08 | 1.57 | 4.68E-03 |
| Body Mass Index | HNF1B | -0.70 | -2.16 | 0.75 | 3.41E-01 | 1.08 | 0.40 | 2.91 | 8.82E-01 |
| Body Mass Index | HNF4A | -0.63 | -1.88 | 0.61 | 3.17E-01 | 1.53 | 1.09 | 2.16 | 1.46E-02 |
| Fasting Glucose | HNF1A | -0.60 | -1.28 | 0.08 | 8.46E-02 | 1.12 | 0.91 | 1.37 | 3.01E-01 |
| Fasting Glucose | HNF1B | -0.51 | -2.64 | 1.62 | 6.37E-01 | 1.26 | 0.30 | 5.21 | 7.52E-01 |
| Fasting Glucose | HNF4A | 1.35 | -0.03 | 2.73 | 5.58E-02 | 0.94 | 0.65 | 1.36 | 7.27E-01 |
| HbA1c | HNF1A | 0.36 | -0.25 | 0.98 | 2.48E-01 | 0.94 | 0.78 | 1.14 | 5.34E-01 |
| HbA1c | HNF1B | -0.48 | -2.20 | 1.24 | 5.82E-01 | 1.62 | 0.56 | 4.69 | 3.69E-01 |
| HbA1c | HNF4A | -0.30 | -1.63 | 1.03 | 6.57E-01 | 1.24 | 0.86 | 1.79 | 2.58E-01 |
| Fasting Insulin | HNF1A | 0.23 | -0.38 | 0.85 | 4.55E-01 | 1.09 | 0.90 | 1.31 | 3.96E-01 |
| Fasting Insulin | HNF1B | -0.44 | -2.29 | 1.41 | 6.42E-01 | 2.23 | 0.73 | 6.82 | 1.58E-01 |
| Fasting Insulin | HNF4A | -0.68 | -2.04 | 0.69 | 3.29E-01 | 0.87 | 0.58 | 1.29 | 4.84E-01 |
| Lipodystrophy | HNF1A | 0.28 | -0.35 | 0.91 | 3.81E-01 | 0.87 | 0.72 | 1.05 | 1.35E-01 |
| Lipodystrophy | HNF1B | 1.59 | -0.25 | 3.43 | 9.06E-02 | 0.46 | 0.13 | 1.58 | 2.15E-01 |
| Lipodystrophy | HNF4A | 1.69 | 0.38 | 3.00 | 1.18E-02 | 1.12 | 0.79 | 1.59 | 5.23E-01 |
| Waist Hip Ratio | HNF1A | -0.52 | -1.13 | 0.10 | 1.01E-01 | 1.00 | 0.83 | 1.21 | 9.72E-01 |
| Waist Hip Ratio | HNF1B | 0.44 | -1.16 | 2.04 | 5.85E-01 | 2.75 | 0.90 | 8.43 | 7.64E-02 |
| Waist Hip Ratio | HNF4A | -0.36 | -1.61 | 0.90 | 5.74E-01 | 0.96 | 0.68 | 1.36 | 8.16E-01 |

Table S6. Effect of Polygenic Score on Phenotype, by MODY subtype. Association results between age of diagnosis (beta) / diabetes severity (odds ratio), split by HNF MODY subtype. Estimates were derived using a mixed-effects linear or logistic model with family as a random effect and adjusted for other polygenic scores and the first ten within-cohort principal components

| Gene | Transcript | dna_nomenclature | protein_nomenclature |
|---|---|---|---|
| *HNF1A* | NM_000545.6 | c.1136C>G | p.Pro379Arg |
| *HNF1A* | NM_000545.6 | c.1309+1G>A | p.? |
| *HNF1A* | NM_000545.6 | c.1330_1331del | p.Gln444Glufs*104 |
| *HNF1A* | NM_000545.6 | c.1396C>T | p.Gln466* |
| *HNF1A* | NM_000545.6 | c.1475C>T | p.Thr492Ile |
| *HNF1A* | NM_000545.6 | c.1487_1494del | p.Leu496Profs*50 |
| *HNF1A* | NM_000545.6 | c.160C>T | p.Arg54* |
| *HNF1A* | NM_000545.6 | c.343G>T | p.Val115Leu |
| *HNF1A* | NM_000545.6 | c.347C>T | p.Ala116Val |
| *HNF1A* | NM_000545.6 | c.391C>T | p.Arg131Trp |
| *HNF1A* | NM_000545.6 | c.392G>A | p.Arg131Gln |
| *HNF1A* | NM_000545.6 | c.404del | p.Asp135Valfs*20 |
| *HNF1A* | NM_000545.6 | c.431T>C | p.Leu144Pro |
| *HNF1A* | NM_000545.6 | c.475C>T | p.Arg159Trp |
| *HNF1A* | NM_000545.6 | c.526C>T | p.Gln176* |
| *HNF1A* | NM_000545.6 | c.527-1G>A | p.? |
| *HNF1A* | NM_000545.6 | c.591G>T | p.Lys197Asn |
| *HNF1A* | NM_000545.6 | c.598C>T | p.Arg200Trp |
| *HNF1A* | NM_000545.6 | c.599G>A | p.Arg200Gln |
| *HNF1A* | NM_000545.6 | c.608G>A | p.Arg203His |
| *HNF1A* | NM_000545.6 | c.646C>T | p.Gln216* |
| *HNF1A* | NM_000545.6 | c.685C>T | p.Arg229* |
| *HNF1A* | NM_000545.6 | c.686G>A | p.Arg229Gln |
| *HNF1A* | NM_000545.6 | c.812G>A | p.Arg271Gln |
| *HNF1A* | NM_000545.6 | c.824_826del | p.Glu275del |
| *HNF1B* | NM_000458.4 | c.1040dup | p.Ser348Valfs*12 |

| | | | |
|---|---|---|---|
| *HNF1B* | NM_000458.4 | c.1654-2A>G | p.? |
| *HNF1B* | NM_000458.4 | c.476C>T | p.Pro159Leu |
| *HNF1B* | NM_000458.4 | c.493C>T | p.Arg165Cys |
| *HNF1B* | NM_000458.4 | c.907C>A | p.Arg303Ser |
| *HNF1B* | NM_000458.4 | 17q12 Microdeletion | |
| *HNF4A* | NM_175914.4 | c.1033G>T | p.Asp345Tyr |
| *HNF4A* | NM_175914.4 | c.124G>A | p.Gly42Arg |
| *HNF4A* | NM_175914.4 | c.322G>A | p.Val108Ile |
| *HNF4A* | NM_175914.4 | c.335G>A | p.Arg112Gln |
| *HNF4A* | NM_175914.4 | c.352C>T | p.Arg118* |
| *HNF4A* | NM_175914.4 | c.469A>C | p.Lys157Gln |
| *HNF4A* | NM_175914.4 | c.530T>C | p.Val177Ala |
| *HNF4A* | NM_175914.4 | c.537G>A | p.Trp179* |
| *HNF4A* | NM_175914.4 | c.614A>C | p.His205Pro |
| *HNF4A* | NM_175914.4 | c.625G>A | p.Gly209Arg |
| *HNF4A* | NM_175914.4 | c.691C>T | p.Arg231Trp |
| *HNF4A* | NM_175914.4 | c.733C>T | p.Arg245Cys |
| *HNF4A* | NM_175914.4 | c.734G>A | p.Arg245His |
| *HNF4A* | NM_175914.4 | c.787G>C | p.Glu263Gln |
| *HNF4A* | NM_175914.4 | c.823C>T | p.Pro275Ser |
| *HNF4A* | NM_175914.4 | c.869G>A | p.Arg290His |
| *HNF4A* | NM_175914.4 | c.925C>T | p.Arg309Cys |
| *HNF4A* | NM_175914.4 | c.926G>A | p.Arg309His |
| *HNF4A* | NM_175914.4 | c.932G>A | p.Arg311His |
| *HNF4A* | NM_175914.4 | c.956T>C | p.Leu319Pro |

Table S8. Pathogenic variants of *HNF1A*, *HNF1B* and *HNF4A* identified in the UK Biobank

| Group | Prevalence | h2 (GCTA-GREML-LDMS) | SE (GCTA-GREML-LDMS) | P (GCTA-GREML-LDMS) | h2 (LDAK-REML) | SE (LDAK-REML) | P (LDAK-REML) | h2 (LDAK-PCGC) | SE (LDAK-PCGC) | P (LDAK-PCGC) |
|---|---|---|---|---|---|---|---|---|---|---|
| All HNF MODY | 0.0005 | 0.239224 | 0.034465 | 1.50E-06 | 0.228802 | 0.033119 | 6.45E-14 | 0.220527 | 0.10972 | 5.41E-03 |
| All HNF MODY | 0.00025 | 0.217268 | 0.031302 | 1.50E-06 | 0.207506 | 0.030037 | 6.45E-14 | 0.202287 | 0.101634 | 5.94E-03 |
| HNF1A MODY | 0.0005 | 0.270959 | 0.052318 | 1.28E-04 | 0.255482 | 0.050068 | 4.29E-08 | 0.321433 | 0.191599 | 1.63E-02 |
| HNF1A MODY | 0.00025 | 0.24609 | 0.047516 | 1.28E-04 | 0.231703 | 0.045408 | 4.29E-08 | 0.29589 | 0.177519 | 1.72E-02 |
| T2D | 0.1 | 0.30841 | 0.029422 | 1.47E-07 | 0.290519 | 0.028217 | 9.81E-28 | 0.278013 | 0.042226 | 4.36E-26 |

Table S9. SNP based heritability estimates for MODY and T2D. Common variant heritability (h2) was estimated on the liability scale (i.e. across various disease prevalences). Estimates were obtained using GCTA-GREML, with LDAK-REML and LDAK-PCGC for sensitivity analysis, and also sub setting to carriers of *HNF1A* MODY. Heritability analysis was restricted to unrelated individuals.
